# Detection of prevalent SARS-CoV-2 variant lineages in wastewater and clinical sequences from cities in Québec, Canada

**DOI:** 10.1101/2022.02.01.22270170

**Authors:** Arnaud N’Guessan, Alexandra Tsitouras, Fernando Sanchez-Quete, Eyerusalem Goitom, Sarah J. Reiling, Jose Hector Galvez, Thanh Luan Nguyen, Ha Thanh Loan Nguyen, Flavia Visentin, Mounia Hachad, Kateryna Krylova, Sara Matthews, Susanne A. Kraemer, Paul Stretenowich, Mathieu Bourgey, Haig Djambazian, Shu-Huang Chen, Anne-Marie Roy, Brent Brookes, Sally Lee, Marie-Michelle Simon, Thomas Maere, Peter A. Vanrolleghem, Marc-Andre Labelle, Sandrine Moreira, Inès Levade, Guillaume Bourque, Jiannis Ragoussis, Sarah Dorner, Dominic Frigon, B. Jesse Shapiro

**Author notes:** these authors jointly supervised this work (Jiannis Ragoussis, Sarah Dorner, Dominic Frigon, B. Jesse Shapiro).

## Abstract

Wastewater-based epidemiology has emerged as a promising tool to monitor pathogens in a population, particularly when clinical diagnostic capacities become overwhelmed. During the ongoing COVID-19 pandemic caused by Severe Acute Respiratory Syndrome Coronavirus-2 (SARS-CoV-2), several jurisdictions have tracked viral concentrations in wastewater to inform public health authorities. While some studies have also sequenced SARS-CoV-2 genomes from wastewater, there have been relatively few direct comparisons between viral genetic diversity in wastewater and matched clinical samples from the same region and time period. Here we report sequencing and inference of SARS-CoV-2 mutations and variant lineages (including variants of concern) in 936 wastewater samples and thousands of matched clinical sequences collected between March 2020 and July 2021 in the cities of Montreal, Quebec City, and Laval, representing almost half the population of the Canadian province of Quebec. We benchmarked our sequencing and variant-calling methods on known viral genome sequences to establish thresholds for inferring variants in wastewater with confidence. We found that variant frequency estimates in wastewater and clinical samples are correlated over time in each city, with similar dates of first detection. Across all variant lineages, wastewater detection is more concordant with targeted outbreak sequencing than with semi-random clinical swab sampling. Most variants were first observed in clinical and outbreak data due to higher sequencing rate. However, wastewater sequencing is highly efficient, detecting more variants for a given sampling effort. This shows the potential for wastewater sequencing to provide useful public health data, especially at places or times when sufficient clinical sampling is infrequent or infeasible.

## INTRODUCTION

Quebec has been one of the Canadian provinces most affected by the COVID-19 pandemic, with 5,698 cases and 135 deaths per 100,000 inhabitants as of December 2021 (Public Health Agency of Canada 2021). After the first reported COVID-19 cases at the end of February 2020, the government implemented public health measures to control the spread of the virus, ranging from the closure of non-essential businesses to intra-provincial travel restrictions. The Coronavirus Sequencing in Quebec (CoVSeQ) consortium began sequencing SARS-CoV-2 genomes from qPCR-positive nasal swabs sampled across the province, yielding insights into the early introduction events of the virus into the province, and their subsequent spread (Murall et al. 2021). As in many other places around the world, the Quebec Public Health lab (LSPQ) increased genomic surveillance of SARS-CoV-2 after the emergence of variants of concern (VOCs) at the end of 2020. Variants of interest (VOIs) are defined as SARS-CoV-2 lineages with mutations that have a potential impact on the clinical or epidemiological characteristics of the virus, while those with a demonstrated impact on disease transmission or clinical features are named VOCs (Institut national de santé publique du Québec 2021). For example, the Alpha variant (PANGO lineage B.1.1.7) had a significant transmission advantage relative to previously circulating lineages (Volz et al. 2021; Davies et al. 2021), which was then superseded by the even more transmissible Delta (B.1.617.2) variant (Campbell et al. 2021; Mlcochova et al. 2021). Although Quebec has the highest testing rate in Canada (491 tests performed per 100,000 habitants per week vs 335 on average in Canada as of December 18, 2021) and a decreasing death rate since the beginning of 2021 (from 4.02 new deaths per 100,000 habitants per week at the beginning of 2021 to 0.0 in mid August 2021), the recent increase in the case incidence rate (from 1.1 new cases per 100,000 habitants in mid July 2021 to 3.3 in mid August 2021) and the threat of new variants with immune escape or further transmission advantages (Otto et al. 2021) calls for continued vigilance and improved surveillance.

Wastewater (WW) surveillance has emerged as a method to track SARS-CoV-2 variants in a manner that is complementary to sequencing clinical samples. Wastewater-based epidemiology has the potential to variant lineages earlier than clinical sampling and provides valuable insights about the viral spread in the population (Bibby et al. 2021; Xiao et al. 2021). This is because wastewater sampling can catch asymptomatic cases that would otherwise not present for a nasal swab sample, but that do excrete viral RNA in stool (Bibby et al. 2021; Jones et al. 2020). Wastewater is a complex mixture of fragmented viral RNA, which can be difficult to sequence and confidently identify mutations. Recent studies in the United States have shown that the dominant SARS-CoV-2 variants observed in clinical samples are largely mirrored in wastewater sequences (Crits-Christoph et al. 2021; Baaijens et al. 2021). Wastewater also has the potential to capture mutations and variant lineages not observed in human clinical samples, potentially including animal reservoirs (Smyth et al. 2021). However, more studies are required to assess the sensitivity and spatio-temporal resolution of wastewater sequencing in comparison to clinical sequencing.

## RESULTS AND DISCUSSION

### Wastewater and clinical SARS-CoV-2 sampling and sequencing in Quebec

As a demonstration of WW-based VOCs and VOIs surveillance, we deep-sequenced SARS-CoV-2 genome-wide amplicons from 936 WW samples collected between March 2020 and July 2021 in the cities of Montreal, Quebec, and Laval, representing almost half of Quebec’s population. The WW samples were collected from interceptors at the end of the WW network (36.1%), residential areas (29.9%), WW treatment facilities (21.2%), prisons (7.69%), industrial zones (4.90%) and long-term care and housing centers (0.21%). We compare the WW data to semi-random clinical samples from the same cities, which we refer to as the clinical dataset (*N* = 13,296 sequences), and non-random priority sequencing of outbreaks and other samples of particular public health interest (*e*.*g*. suspected Alpha variant cases in early 2021, travel-related cases, etc.), which we refer to as ‘outbreak’ samples (*N* = 5,661 sequences).

### Establishing thresholds for calling single nucleotide variants (SNVs) with confidence

To study the genomic diversity of SARS-CoV-2 in WW samples, we first developed a bioinformatic pipeline to confidently identify single nucleotide variants (SNVs) while accounting for sequencing errors, then to infer VOIs, VOCs, and other variant lineages that contain combinations of signature SNVs (**Methods**). WW samples can contain a mixture of variants at different frequencies. True SNVs can be confused with errors that are introduced by sample processing, e.g. from errors introduced by the reverse transcriptase or the polymerase that are used for targeted SARS-CoV-2 amplification, or from read errors created by the sequencing platform. In this study, we chose illumina sequencing to minimize the per-read error rate (compared to Oxford Nanopore). To set appropriate thresholds for minimum SNV frequencies and coverage, we sequenced 14 SARS-CoV-2 positive control genomes from AccuGenomics, containing a set of known SNVs at 100% within-sample frequency, which we sequenced using the same Illumina amplicon strategy used for the WW samples. We assessed a range of filters for depth of coverage and minor allele frequency (MAF) required to call a SNV (**Figure 1**). Our SNV calling pipeline yields systematically low false positive rates (median < 7% even at 1% MAF and effectively zero at 25% MAF) across a range of filters and expected frequency of the true positive SNVs (**Figure 1**). To be conservative for subsequent analyses we selected a minimum site coverage of 50X and a MAF of 25%. These filters maximized the F1 score, which combines information about the precision (i.e. the proportion of SNV calls that are true positives) and the sensitivity (i.e. the proportion of true variants that are identified). The F1 scores are poor when the expected frequency of the true positive SNVs is 100% (**Figure S1A**) but improves when we allow the expected nucleotide to have a frequency of at least 75% to allow for sequencing errors (**Figure S1B**). Note that very similar F1 scores were obtained at 10% MAF and that our variant calling pipeline can detect low-frequency mutations with high accuracy (**Figures 1 and S2**), but we proceeded with 25% to be conservative. In addition to the AccuGenomics controls, we also sequenced mixtures of two different SARS-CoV-2 viral cultures at known ratios, which we call “spike-in” samples. We found that our SNV-calling pipeline identified the expected SNVs close to their expected frequencies. Low variant frequencies (expected frequency < 5%) were inferred particularly accurately, with more variation in the 25-50% range (**Figure S2**). Overall, these results indicate that our SNV-calling filters are appropriate, and likely conservative, for identifying SNVs in wastewater containing mixtures of viral genomes.

**Figure 1.**
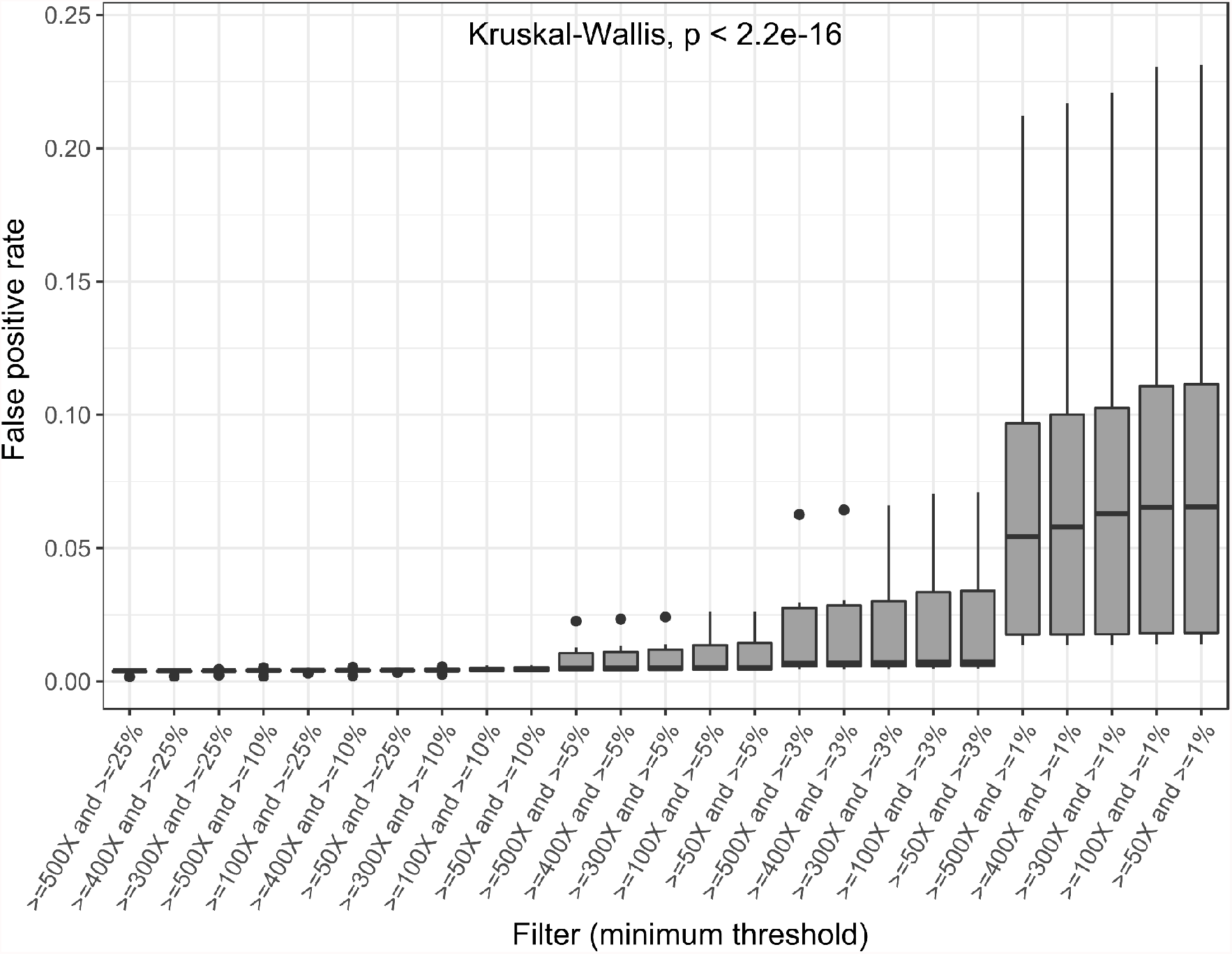
False-positive SNV calling rates across different depth and frequency filters. In all cases, the expected variant frequency was 100% in AccuGenomics SARS-CoV-2 genome standards. The thresholds that define the filters are respectively the minimum coverage (X) and the minimum minor SNV frequency. The Kruskal-Wallis test p-value (at the top of the panel) indicates the significance of the differences across the different sets of filters.

### Sampling and sequencing SARS-CoV-2 from wastewater

Having established a reliable method for SNV calling, we applied it to a set of 936 WW samples collected between March 2020 and July 2021 in Montreal, Quebec City, and Laval (**Table S1**). Because SNV calling requires sufficient depth of coverage, we measured how coverage depth and breadth varied across the SARS-CoV-2 genome in our samples (**Figure 2**).

**Figure 2.**
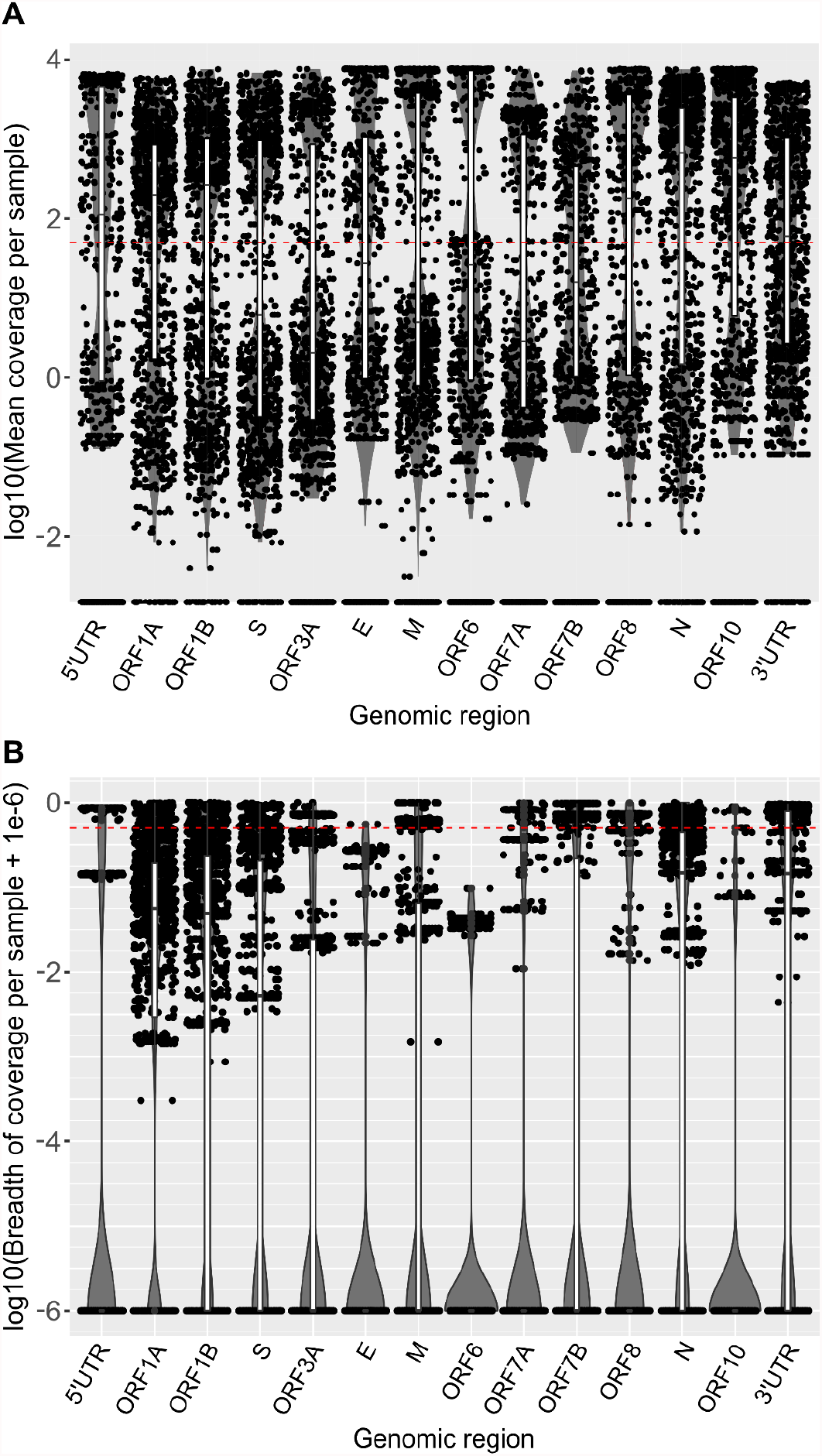
Sequencing coverage across the SARS-CoV-2 genome recovered from wastewater. A) Depth of coverage, defined as the average number of sequencing reads covering a nucleotide site. Each point represents a unique wastewater sample. Each distribution is represented by a jitter plot (black points), a boxplot (black and white) and a violin plot (grey). The red dotted line indicates a coverage of 50 on a log10 scale. B) Breadth of coverage. The breadth of coverage represents the proportion of sites in each genomic region (gene or open reading frame; ORF) with a coverage of at least 50X. Boxplots are absent when 75% of the samples have a breadth of coverage of 0 or, in other words, the 3 first quantiles of the distribution correspond to samples with a coverage <50 at the corresponding sites. The red dotted line indicates a breadth of coverage of 50% on a log10 scale.

The mean depth and breadth of coverage varied somewhat across genes, and was roughly bimodal, with samples tending to fall either close to 0-1X coverage or >1000X on average, with fewer samples in between (**Figure 2**). As expected, coverage correlated negatively with PCR Cycle threshold (linear regression adjusted R^2^ = 19.2%; Permutational ANOVA *p* < 2e-4, n = 5000 permutations), as previously observed (Lythgoe et al. 2021; Baaijens et al. 2021). Due to this variation in coverage, not all nucleotide sites in the genome in all samples have sufficient depth to make SNV calls. However, given the substantial number of samples with >1000X coverage, we expect to obtain a reasonable number of high-confidence SNVs in our dataset. Using the >=50X coverage and 25% MAF filters described above, we called an average of 6.87 SNVs per sample (s.d. = 10.1) (**Figure S3**).

### Detecting SARS-CoV-2 lineages in wastewater using signature and marker mutations

With these SNVs in hand, we sought to infer the presence of known variant lineages in WW samples. We defined “signature mutations” as SNVs with a minimum prevalence of 90% among the consensus sequences of a certain lineage (*e*.*g*. a particular VOC represented in a database). To find these signature mutations, we first calculated the prevalence of substitutions in thousands of publicly available consensus sequences collected during 2020 and added data from CoV-Spectrum (Chen et al. 2021) to include under-represented lineages or lineages that emerged after 2020 (**Methods**). These variant lineages were named using the PANGO lineage designation scheme (Rambaut et al. 2020). We further defined “marker mutations” as signature mutations that are unique to a single variant lineage to the exclusion of others.

Using these criteria, we identified an average of 4.42 signature mutations per sample (s.d. = 6.94) and 2.01 marker mutations per sample (s.d. = 3.22) (**Figure S3**). To call a variant lineage as present in a WW sample, we required at least 3 signature mutations including at least one marker mutation (**Methods**), yielding an average of 0.79 variant lineages identified per sample (s.d. = 1.14; min.=0; max. = 6). This approach allowed us to track the spread of variants with a confidence score, i.e. the number of signature mutations supporting the presence of a lineage, which includes at least one of its marker mutations (**Figure S4 and Table S2**). As expected in this time period, the samples are dominated by the Alpha variant (B.1.1.7), with sporadic identification of other variants.

We also estimated the within-sample frequency of variant lineages using constrained linear models (**Methods**). These models infer the linear combination of known variant lineages (from the database of reference genomes) that best explain the frequencies of observed SNVs in the sample (**Figure S6**). We applied this approach to the 299 WW samples that have at least one lineage with at least 3 signature mutations, including at least one marker mutation. Among these samples, the regression model was significant for 250 samples (83.6%) and not significant for the remaining 49 samples (16.4%). These variant frequency estimates (**Figure S5**) were generally concordant with the simpler approach relying on signature and marker mutations (**Figure S4**). Both approaches showed a predominance of the Alpha variant in all cities, with substantial numbers of B.1.160 in 2020 and early 2021, and A.2.5.X lineages previously described in Quebec clinical samples (Murall et al., 2021).

The absence of variant lineage detections can simply be due to lack of sampling, lack of sequencing depth, or a true absence. Sampling frequency is indeed variable over time and cities (**Figure S7**) and the absence of detections is mostly explained by the absence of WW samples (81.4% of time points with missing lineages; grey shading in **Figures S4 and S5**) and modestly by lack of sufficient sequencing depth (18.6%; transparent red shading in **Figures S4 and S5**). This suggests that increased sampling frequency, coupled with modest optimization of RNA extraction and sequencing protocols, could increase both the true positive and true negative rates.

### Comparisons between wastewater and nasal swab sequencing

To assess how SARS-CoV-2 variant lineage detection in wastewater compared with sequencing of clinical samples in the same cities and time period, we considered the top three most prevalent lineages detected in our WW samples: Alpha (B.1.1.7), B.1.160, and A.2.5.X). For these analyses, we supplemented the 936 WW sequences with 13,296 clinical samples from a semi-random population sample of nasal swabs (of which 9,262 passed sequencing quality filters and were deposited in GISAID; **Table S3**) and 5,661 non-random ‘outbreak’ samples sequenced with high priority by the Quebec Public Health lab (of which 1,848 passed filters and were deposited in GISAID; **Table S4**; Methods). In each sample type, we defined a variant lineage’s frequency relative to other lineages in a 2-week time window. The frequency estimates from WW and both types of clinical samples are significantly correlated (linear regression adjusted R^2^ = 0.63 with semi-random clinical data, and 0.34 with outbreak data; Permutational ANOVA both *p* < 2e-4, pooled over the three most prevalent variants; n = 5000 permutations). Note that this method ignores missing data, so sampling gaps do not contribute to the regression. In general, variants are detected first in clinical or outbreak data, and shortly thereafter in wastewater (**Figure 3**). The Alpha VOC (B.1.1.7) in particular tended to be detected first in outbreak samples, likely because suspected Alpha cases were prioritized for sequencing by the Quebec Public Health lab at the time. Notably, Alpha was detected in WW concurrently or shortly after outbreak samples, despite being detected much later (or not at all) in semi-random clinical sequences (**Figure 3**).

**Figure 3.**
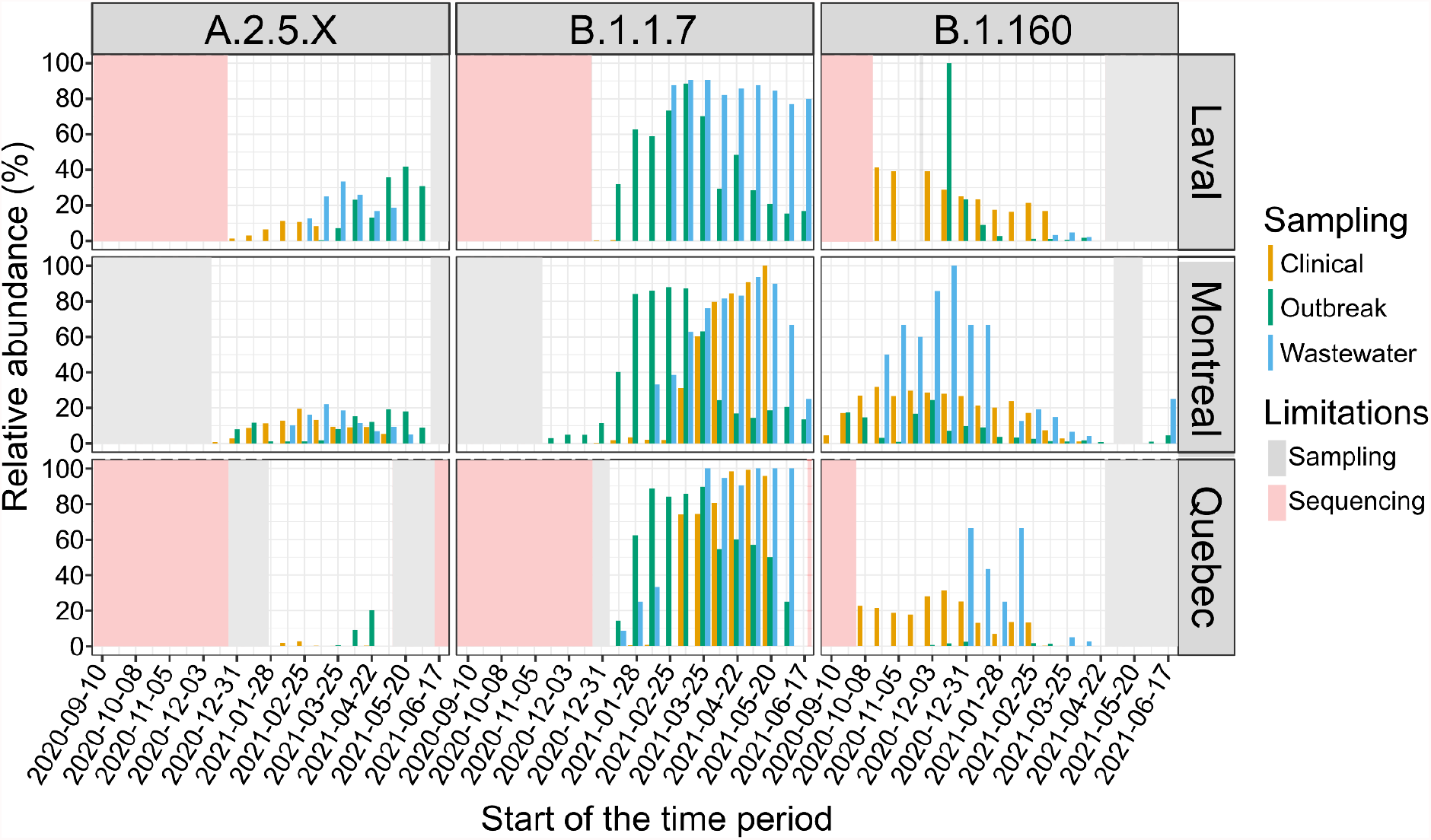
Relative abundance of the most prevalent VOC/VOIs in Quebec in clinical, outbreak and WW samples. The relative abundance of each variant was estimated as the percentage of samples in which the variant is present over a 2-week period. Gaps in the time series can be explained by a lack of sampling (transparent grey), i.e. the absence of detections of a particular lineage due to the absence of samples, or missing detections in the sequencing data (transparent red), i.e. the absence of detections of a particular lineage although samples were collected during the corresponding period. We only show time intervals starting in September 2020 because sampling frequency was sparse before that (Figure S7).

Next, we sought to generalize these results beyond the most prevalent variants and to consider different possible time-lags between clinical and WW samples. For example, clinically undetected asymptomatic or pre-symptomatic cases could be detected earlier with WW sampling. Conversely, outbreak and clinical samplings might detect some lineages earlier than WW when the testing rate is high and appropriately targeted. We considered time lags between 0 and 8 days between collection of WW and clinical or outbreak samples, and calculated concordance, defined simply as the detection of a variant in both sample types. We found that concordance was maximized between WW and both clinical or outbreak samples at a maximum time lag of around 7 days, regardless of the number of signature SNVs required to define variants (**Figures S8 and S9**). The 7-day time interval is similar to the average pre-symptomatic phase duration, i.e. 6.4 days (Backer, Klinkenberg, and Wallinga 2020). To assess the significance of these concordance scores, we permuted (1000 times) the variant detections across cities and time (days) and compared the concordance score after permutations to the original concordance score to obtain a *p*-value. Using a 7-day maximum time lag and defining variants with at least three signature SNVs, we estimated that 41.7% of WW calls are concordant with semi-random clinical sampling data (permutation test *p*-value = 0.23) and 85.7% of WW calls are concordant with outbreak sampling data (permutation test *p*-value = 0.009). None of the tested time gaps yield a significant concordance between WW and semi-random clinical data (**Figure S8**). The stronger concordance between WW and outbreak sequences is surprising, as one would expect WW to better capture the same variants as the semi-random clinical samples. Although the reasons for this observation are unclear, it speaks to the unknown and potentially orthogonal biases implicit in each of the sampling schemes. Out of the 14 variant lineages detected both in WW and outbreak datasets, 13 are also detected in the clinical dataset (**Figure S10**). This suggests that the weaker concordance between WW and clinical data relative to outbreak data is not explained by the identity of the lineage in question, but rather by discrepancies in the timing of detection. Indeed, 37 lineages were detected earlier by semi-random clinical sampling than WW or outbreak sampling, including A.2.5.X, B.1.1.7 (Alpha), B.1.160, B.1.526 (Iota) and R.1, while seven lineages were detected earlier in the outbreak dataset, including P.1 (Gamma), B.1.617.X (Kappa/Delta), B.1.1.519 and C.37 (Lambda). Only B.1.351 (Beta), B.1.621 (Mu) and B.1.214.2 were detected first in the WW data.

A major reason that more variant lineages are detected earlier in clinical and outbreak sequences is simply the sampling effort: there are about five times more available outbreak samples than WW samples in our dataset, and about ten times more semi-random clinical samples. As a result, there are more lineage detections in clinical data (**Figure S11A**), which is consistent with another study from the United States (Baaijens et al. 2021) and is explained by a higher monthly sequencing rate (**Figure S11B**). However, at an equal sequencing rate, WW is able to detect more variant lineages than clinical or outbreak sampling (**Figure 4**). This suggests that, for a given sequencing effort, WW surveillance could detect a higher diversity of variants. Even in our current limited sample of 936 WW sequences, we are able to infer the presence of certain variants that are not apparent in clinical sequences (**Figure S10**).

**Figure 4.**
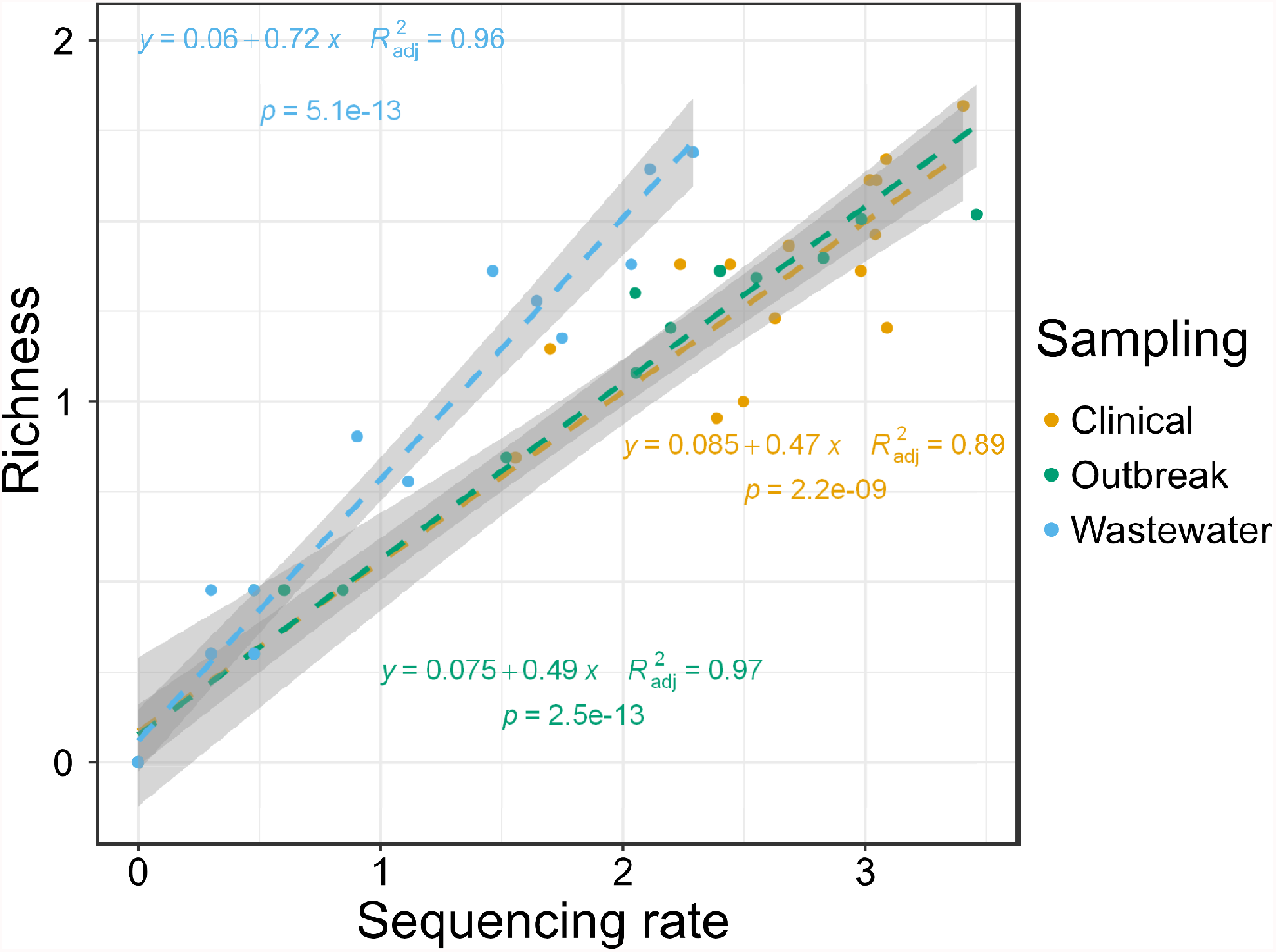
Wastewater sampling detects more unique variant lineages at a given sequencing rate. Monthly richness of detected lineages in function of sequencing rate for clinical (gold), outbreak (green) and wastewater (skyblue) samplings. We estimated the sequencing rate as *log10(number of sequences per month+1)* and richness as *log10(number of variant lineages detected +1)*.

## CONCLUSION

Using a relatively conservative SNV calling pipeline and lineage detection method, we tracked the spread of SARS-CoV-2 lineages in wastewater between March 2020 and July 2021 across three cities in the province of Quebec. Although WW sequencing could be used to detect novel mutations or variant lineages not found in clinical samples (Smyth et al. 2021), here we focused on identifying known variants. Consistent with a similar study in the US, we found that sequencing coverage in WW was dependent on viral load, as measured by the qPCR cycle threshold (Ct) value (Baaijens et al. 2021). This suggests that WW sample concentration or optimization of RNA extraction could yield more sequence data and improve variant inference. We also found that the most prevalent variants were detected concordantly in WW and clinical samples over a 7-day time frame, but that variants were generally detected first in clinical samples, suggesting that clinical diagnostics efforts were effective at the time of sampling. We note that certain VOCs and VOIs, including B.1.351 (Beta), B.1.214.2, and B.1.621 (Mu) were detected in WW before clinical samples. Several other variants were detected only in WW but not clinical samples. These could be true positives that were undetected by clinical sampling, or false positives that could have arisen for a variety of reasons, including the threshold number of signature or marker SNVs required to identify a lineage. In other words, even if we trust the SNV calls, recurrent SNVs could potentially confuse one variant lineage for another. Outbreak samples sequenced as part of targeted public health investigations were particularly concordant with WW samples, for reasons that remain unclear. Despite sources of error in sampling and sequencing, WW-based detection of variant lineages is generally aligned with clinical samples from the same city and time period. Our analyses pooled a variety of sample types at the city level. Future studies could examine the spatial scales (province, city, neighborhood, residence) at which wastewater sequencing is most concordant with clinical sampling.

Importantly, WW samples are able to detect more variant lineages per sequencing run. This is intuitive, because WW includes a mixture of viruses, thereby often sampling multiple individuals at a time, depending on the wastewater catchment area. This mixture of viruses must then be inferred based on sequencing data. Here we used Illumina short-read data, and a SNV calling pipeline benchmarked on known standard SARS-CoV-2 genomes. We then inferred variant lineages as combinations of SNVs present in known variants, based on either a simple threshold of signature and marker mutations, or on a constrained linear model, yielding similar results. Long-read technology is potentially useful for resolving multiple SNVs on the same sequencing read, potentially providing more direct evidence for variant genomes and circumventing the need for inference based on unlinked SNV frequencies. However, RNA in wastewater is generally quite fragmented, which may impose an upper limit on the utility of such methods. Furthermore, the single read accuracy of Nanopore sequencing is to date not at par with the read accuracy offered by Illumina.

While there is room for methodological improvements ranging from sample collection, sequencing, and computational inference of variants, the general concordance between wastewater and clinical sampling is encouraging. In contexts where clinical sampling is infrequent or infeasible, wastewater can provide a complementary window into VOC frequencies. The value of wastewater sequencing became exceedingly clear during the Omicron wave, which began in Canada in mid-December 2021 and is still ongoing in late January 2022. Anecdotally, we were able to detect Omicron with 11 signature mutations on December 4, 2021 in Montreal (data not shown). Together, our results suggest that wastewater sequencing can continue to provide a similar portrait of SARS-CoV-2 variant lineages, potentially with much less sampling and sequencing effort.

## Data Availability

Raw wastewater sequencing data is available in the NCBI SRA database under BioProject ID PRJNA788395 (http://www.ncbi.nlm.nih.gov/bioproject/788395O). Viral genomes from clinical samples are available in GISAID under IDs listed in Tables S3 and S4. The SNV calling and post-variant-calling pipeline is available at: https://github.com/arnaud00013/Wastewater_surveillance_pipeline

https://www.gisaid.org/

## ACKNOWLEDGEMENTS

We are particularly grateful to Carole Fleury, Suzanne Boulet, Luc Carrière from the City of Montreal, David Kaiser from Montreal Public Health, Mario Gagné from the City of Laval and Diarra Zeinab from the Centre de technologies de l’eau for their assistance in obtaining samples. We also thank Frédéric Cloutier, Denis Dufour and François Proulx from Quebec City for assistance with sampling and logistics support.

This study was supported by the Canadian Institutes for Health Research (CIHR) operating grant to the Coronavirus Variants Rapid Response Network (CoVaRR-Net) to JR and BJS and the CFI project “High throughput SARS-CoV-2 genome sequencing at the McGill Genome Center” to JR. DF was supported by the Fonds de Recherche du Québec (FRQ) and a McGill MI4 Emergency COVID-19 Research Fund grant. SD was supported by FRQ and The Trottier and Molson Foundations. Data analyses were enabled by compute and storage resources provided by Compute Canada and Calcul Québec. We would like to acknowledge the team at the Canadian Centre for Computational Genomics for their contributions to the development and testing of the pipeline used for these analyses.

## ETHICS STATEMENT

This investigation was carried out in accordance with the legal mandate granted to public health authorities by the Public Health Act (LRQ, chapter S-2.2. Article 1; http://legisquebec.gouv.qc.ca/fr/ShowDoc/cs/S-2.2) as part of a public health intervention. All data was treated confidentially and analysed without nominal identification in accordance with the Policy on Information Protection and Security (PO-04-2014) of the National Public Health Institute of Quebec (INSPQ). The INSPQ therefore deemed this study exempt from ethical oversight according to provincial legislation.

## DATA & CODE AVAILABILITY

Raw wastewater sequencing data is available in the NCBI SRA database under BioProject ID PRJNA788395 (http://www.ncbi.nlm.nih.gov/bioproject/788395). Viral genomes from clinical samples are available in GISAID under IDs listed in Tables S3 and S4. The SNV calling and post-variant-calling pipeline is available at: https://github.com/arnaud00013/Wastewater_surveillance_pipeline.

## METHODS

### Sample collection and sequencing

To perform the genomic surveillance of SARS-CoV-2, we collected WW samples at the institutional, district, and municipal scale from Montreal, Quebec City, and Laval. The samples were collected by composite sampling, grab sampling, and passive sampling. The composite samples were collected with autosamplers, which collected wastewater every 10 minutes, over a 24, 48, or 72 hour time period. The passive samples were collected through 2 absorbent materials, Q-tips and negatively charged membranes, Mixed Cellulose Ester (MCE) filters, which were housed in torpedoes (Schang et al. 2021). The torpedoes were also deployed over a 24, 48, or 72 hour time period. For time series analyses, the samples were pooled from the different scales (institution, district) and from the different sampling methods (composite, grab, and passive) for each date; thus the analyses were conducted at the municipal scale. Wastewater samples, grab or composite, were additionally concentrated by filtration. First, in 50 ml of wastewater, the pH was adjusted to between 3.5-4.5, and MgCl was added to a final concentration of 25 mM. Then the samples were filtered through a 0.45 μm MCE filter. All samples were processed within at most 72 hours; the MCE filters and Q-tips were stored at - 80°C. RNA was then extracted using the Allprep Powerviral DNA/RNA kit. The protocol was followed according to the manufacturer, with the exception of the lysis step, where a final concentration of 10% Beta-Mercaptoethanol was used in the lysis buffer and incubation time was raised to 30 minutes at 55°C. After extraction, RNA samples were submitted to the McGill Genome Center for reverse transcription followed by targeted SARS-CoV-2 amplification using the ARTIC V3 primer scheme (https://artic.network/resources/ncov/ncov-amplicon-v3.pdf). Samples were purified and a Nextera DNA Flex library preparation was performed for Illumina paired-end amplicon sequencing (PE150) on a NovaSeq instrument at the McGill Genome Center. The detailed protocol can be accessed at: dx.doi.org/10.17504/protocols.io.by6xpzfn.

### SNV calling and quality control

For each sample, we performed quality control of the raw reads using fastp (v.0.20.0, read length >=70, Phred Score >20 and cut_tail option (S. Chen et al. 2018). We then aligned reads to the reference genome (MN908947.3 or NC_045512.2) using bwa (v.0.7.17) (Li 2013) and performed a coverage analysis with samtools (v.1.10) depth (Li 2011). In total, 936 WW samples were collected and sequenced with Illumina. For each of these samples, we performed SNV calling from the read mapping using samtools (v.1.10) mpileup and varscan (v.2.4.1) pileup2snp (Li 2011; Koboldt et al. 2012). To select coverage and SNV frequency filters for SNV calling, we analyzed 14 positive controls from AccuGenomics for which we know the list of expected mutations (https://accugenomics.com/accukit-sars-cov-2/). These controls allow us to measure a limit of detection and to evaluate the background error due to library preparation, sequencing, or other sources of noise. Using these controls, we can then determine the variant calling filters that allows to minimize the background error, i.e. the false positive and false negative rates, and maximizes the accuracy of SNV calling, i.e. F1 score. We calculated this score using the precision, i.e. the proportion of SNV calls (positives) that are true positives, and the recall/sensitivity, i.e. the proportion of expected variants that have been called (Powers 2020). The SNV calling and post-variant-calling pipeline is available at https://github.com/arnaud00013/Wastewater_surveillance_pipeline.

In addition to the AccuGenomics controls, we also sequenced mixtures of two different SARS-CoV-2 viral cultures at known ratios, which we called “spike-in” samples. In these samples, a viral culture extract was spiked into a SARS-CoV-2 positive control sample at a concentration of 1%, 2%, 5%, 10%, 20% or 50%. Both samples were obtained from the first wave of the pandemic and differed by 11 mutations. We performed variant calling on the sequenced spiked-in samples based on the same method as the WW samples, except for the VAF filter, which we removed to be able to detect expected low-frequency mutations. Because each Spike-sample is a mixture of variants and are thus not clonal, the expected variant frequency (VAF) is calculated based on the virus concentration and initial VAF in the samples. *Expected_VAF = (Concentration_positive_control * VAF_in_postive_ctrl) + (Concentration_ViralCulture * VAF_in_L00241026_ ViralCulture)* where *Concentration_positive_control* would be 1%, 2%, 5%, 10%, 20% and 50%, and *Concentration_ViralCulture* = 1 - *Concentration_positive_control*.

### Detection of lineages of interest in WW samples and estimation of their within-sample frequency

To infer the presence of SARS-CoV-2 lineages in WW samples, we used the number of lineage signature mutations, i.e. the number of mutations that have a minimum prevalence of 90% among the consensus sequences of the lineage. To find these signature mutations, we first calculated the prevalence of substitutions in thousands of publicly available consensus sequences collected during 2020 and added data from CoV-Spectrum about under-represented lineage in the database or lineages that emerged during 2021 (C. Chen et al. 2021). The database contains 755 lineages and is available at https://github.com/arnaud00013/Wastewater_surveillance_pipeline/blob/main/Post_variant_calling_analysis/Database_all_mutations_prevalence_in_SC2_lineages_consensus_sequences_as_of_2021_07_08.json. At least 3 signature mutations including a marker mutation, i.e. a substitution that have a very high prevalence (>=90%) only among consensus sequences from a certain lineage, are required to call a lineage present in a WW sample. We selected this filter arbitrarily as a compromise between confidence of detection and sensitivity/stringency. We also made sure to evaluate the effect of selecting different filters on the concordance between WW and clinical sampling.

For the calculation of within-sample frequency of SARS-CoV-2 lineages, we used a linear model to fit the lineages’ signature mutations data to the within-sample mutation frequency data. The rationale of the approach is that the frequency of lineage signature mutations within a sample is a linear combination of the frequency of the lineages and the prevalence of the mutations in the consensus sequences of these lineages. To make sure that linear regression converged to a good solution, we needed to apply certain constraints (**Figure S6**). Thus, we implemented this analysis using the “ConsReg()” function from the R (R Development Core Team 2011) package ConsReg (v.0.1.0), which allows us to perform linear regressions under constraints for regression coefficients. We explored the space of solutions using a Grid Search (coefficients initial values = 0.1,0.5 or 0.9) and a Monte Carlo Markov Chain optimizer.

## REFERENCES

Baaijens, Jasmijn A., Alessandro Zulli, Isabel M. Ott, Mary E. Petrone, Tara Alpert, Joseph R. Fauver, Chaney C. Kalinich, et al. 2021. “Variant Abundance Estimation for SARS-CoV-2 in Wastewater Using RNA-Seq Quantification.” medRxiv, September, 2021.08.31.21262938.

Backer, Jantien A., Don Klinkenberg, and Jacco Wallinga. 2020. “Incubation Period of 2019 Novel Coronavirus (2019-nCoV) Infections among Travellers from Wuhan, China, 20-28 January 2020.” Euro Surveillance 25 (5). https://doi.org/10.2807/1560-7917.ES.2020.25.5.2000062.

Bibby, Kyle, Aaron Bivins, Zhenyu Wu, and Devin North. 2021. “Making Waves: Plausible Lead Time for Wastewater Based Epidemiology as an Early Warning System for COVID-19.” Water Research 202 (September): 117438.

Campbell, Finlay, Brett Archer, Henry Laurenson-Schafer, Yuka Jinnai, Franck Konings, Neale Batra, Boris Pavlin, et al. 2021. “Increased Transmissibility and Global Spread of SARS-CoV-2 Variants of Concern as at June 2021.” Euro Surveillance 26 (24). https://doi.org/10.2807/1560-7917.ES.2021.26.24.2100509.

Chen, Chaoran, Sarah Nadeau, Michael Yared, Philippe Voinov, Ning Xie, Cornelius Roemer, and Tanja Stadler. 2021. “CoV-Spectrum: Analysis of Globally Shared SARS-CoV-2 Data to Identify and Characterize New Variants.” Bioinformatics, December. https://doi.org/10.1093/bioinformatics/btab856.

Chen, Shifu, Yanqing Zhou, Yaru Chen, and Jia Gu. 2018. “Fastp: An Ultra-Fast All-in-One FASTQ Preprocessor.” Bioinformatics 34 (17): i884–90.

Crits-Christoph, Alexander, Rose S. Kantor, Matthew R. Olm, Oscar N. Whitney, Basem Al-Shayeb, Yue C. Lou, Avi Flamholz, et al. 2021. “Genome Sequencing of Sewage Detects Regionally Prevalent SARS-CoV-2 Variants.” mBio 12 (1): e02703–20.

Davies, Nicholas G., Sam Abbott, Rosanna C. Barnard, Christopher I. Jarvis, Adam J. Kucharski, James D. Munday, Carl A. B. Pearson, et al. 2021. “Estimated Transmissibility and Impact of SARS-CoV-2 Lineage B.1.1.7 in England.” Science, March. https://doi.org/10.1126/science.abg3055.

Institut national de santé publique du Québec. 2021. “Définitions Pour La Vigie Sanitaire Des Variants Du SRAS-CoV-2 et Classification Des Lignées Détectées Au Québec.” 2021. https://www.inspq.qc.ca/sites/default/files/publications/3138-definition-vigie-sanitaire-variants-sras-cov-2.pdf.

Jones, David L., Marcos Quintela Baluja, David W. Graham, Alexander Corbishley, James E. McDonald, Shelagh K. Malham, Luke S. Hillary, et al. 2020. “Shedding of SARS-CoV-2 in Feces and Urine and Its Potential Role in Person-to-Person Transmission and the Environment-Based Spread of COVID-19.” Science of the Total Environment 749 (December): 141364.

Koboldt, Daniel C., Qunyuan Zhang, David E. Larson, Dong Shen, Michael D. McLellan, Ling Lin, Christopher A. Miller, Elaine R. Mardis, Li Ding, and Richard K. Wilson. 2012. “VarScan 2: Somatic Mutation and Copy Number Alteration Discovery in Cancer by Exome Sequencing.” Genome Research 22 (3): 568–76.

Li, Heng. 2011. “A Statistical Framework for SNP Calling, Mutation Discovery, Association Mapping and Population Genetical Parameter Estimation from Sequencing Data.” Bioinformatics 27 (21): 2987–93.

Li, Heng. 2013. “Aligning Sequence Reads, Clone Sequences and Assembly Contigs with BWA-MEM.” arXiv [q-bio.GN]. arXiv. http://arxiv.org/abs/1303.3997.

Lythgoe, Katrina A., Matthew Hall, Luca Ferretti, Mariateresa de Cesare, George MacIntyre-Cockett, Amy Trebes, Monique Andersson, et al. 2021. “SARS-CoV-2 within-Host Diversity and Transmission.” Science, March. https://doi.org/10.1126/science.abg0821.

Mlcochova, Petra, Steven A. Kemp, Mahesh Shanker Dhar, Guido Papa, Bo Meng, Isabella A. T. M. Ferreira, Rawlings Datir, et al. 2021. “SARS-CoV-2 B.1.617.2 Delta Variant Replication and Immune Evasion.” Nature, September. https://doi.org/10.1038/s41586-021-03944-y.

Murall Carmen Lía, Eric Fournier, Jose Hector Galvez, Arnaud N’Guessan, Sarah J. Reiling, Pierre-Olivier Quirion, Sana Naderi, et al. 2021. “A Small Number of Early Introductions Seeded Widespread Transmission of SARS-CoV-2 in Québec, Canada.” Genome Medicine 13 (1): 169.

Murall Carmen Lía, Fatima Mostefai, Jean-Christophe Grenier, Raphaël Poujol, Julie Hussin, Sandrine Moreira, B. Jesse Shapiro, CoVSeQ consortium. 2021. “Recent Evolution and International Transmission of SARS-CoV-2 Clade 19B (Pango A Lineages).” June 2, 2021. https://virological.org/t/recent-evolution-and-international-transmission-of-sars-cov-2-clade-19b-pango-a-lineages/711.

Otto, Sarah P., Troy Day, Julien Arino, Caroline Colijn, Jonathan Dushoff, Michael Li, Samir Mechai, et al. 2021. “The Origins and Potential Future of SARS-CoV-2 Variants of Concern in the Evolving COVID-19 Pandemic.” Current Biology. https://doi.org/10.1016/j.cub.2021.06.049.

Powers, David M. W. 2020. “Evaluation: From Precision, Recall and F-Measure to ROC, Informedness, Markedness and Correlation.” arXiv [cs.LG]. arXiv. http://arxiv.org/abs/2010.16061.

Public Health Agency of Canada. 2021. “COVID-19 Daily Epidemiology Update.” 2021. https://health-infobase.canada.ca/covid-19/epidemiological-summary-covid-19-cases.html?stat=rate&measure=deaths&map=pt#a2.

Rambaut, Andrew, Edward C. Holmes, Áine O’Toole, Verity Hill, John T. McCrone, Christopher Ruis, Louis du Plessis, and Oliver G. Pybus. 2020. “A Dynamic Nomenclature Proposal for SARS-CoV-2 Lineages to Assist Genomic Epidemiology.” Nature Microbiology 5 (11): 1403–7.

R Development Core Team. 2011. “R: A Language and Environment for Statistical Computing.” R Foundation for Statistical Computing.

Schang, Christelle, Nicolas D. Crosbie, Monica Nolan, Rachael Poon, Miao Wang, Aaron Jex, Nijoy John, et al. 2021. “Passive Sampling of SARS-CoV-2 for Wastewater Surveillance.” Environmental Science & Technology 55 (15): 10432–41.

Smyth, Davida S., Monica Trujillo, Devon A. Gregory, Kristen Cheung, Anna Gao, Maddie Graham, Yue Guan, et al. 2021. “Tracking Cryptic SARS-CoV-2 Lineages Detected in NYC Wastewater.” medRxiv. https://doi.org/10.1101/2021.07.26.21261142.

Volz, Erik, Swapnil Mishra, Meera Chand, Jeffrey C. Barrett, Robert Johnson, Lily Geidelberg, Wes R. Hinsley, et al. 2021. “Assessing Transmissibility of SARS-CoV-2 Lineage B.1.1.7 in England.” Nature 593 (7858): 266–69.

Xiao, Amy, Fuqing Wu, Mary Bushman, Jianbo Zhang, Maxim Imakaev, Peter R. Chai, Claire Duvallet, et al. 2021. “Metrics to Relate COVID-19 Wastewater Data to Clinical Testing Dynamics.” medRxiv. June. https://doi.org/10.1101/2021.06.10.21258580.

