## Supplementary Figures for "Detection of prevalent SARS-CoV-2 variant lineages in wastewater and clinical sequences from cities in Québec, Canada"

**SUPPLEMENTARY MATERIAL**

**Tables**

**Table S1. List of wastewater sample IDs.**

**Table S2. Lineage detection time series data.** For each lineage present in WW at a certain time and location, the table indicates their marker and signature mutations. The samples that are absent from this list are not published in GISAID because they do not pass the quality filters (due to frameshifts or lower consensus completeness) and are not expected to contribute to the lineage detections.

**Table S3. List of semi-random clinical samples and GISAID IDs.** Sampling dates, PANGO lineages and other meta-data are also included. The samples that are absent from this list are not published in GISAID because they do not pass the quality filters (due to frameshifts or lower consensus completeness) and are not expected to contribute to the lineage detections.

**Table S4. List of outbreak samples and GISAID IDs.** Sampling dates, PANGO lineages and other meta-data are also included.

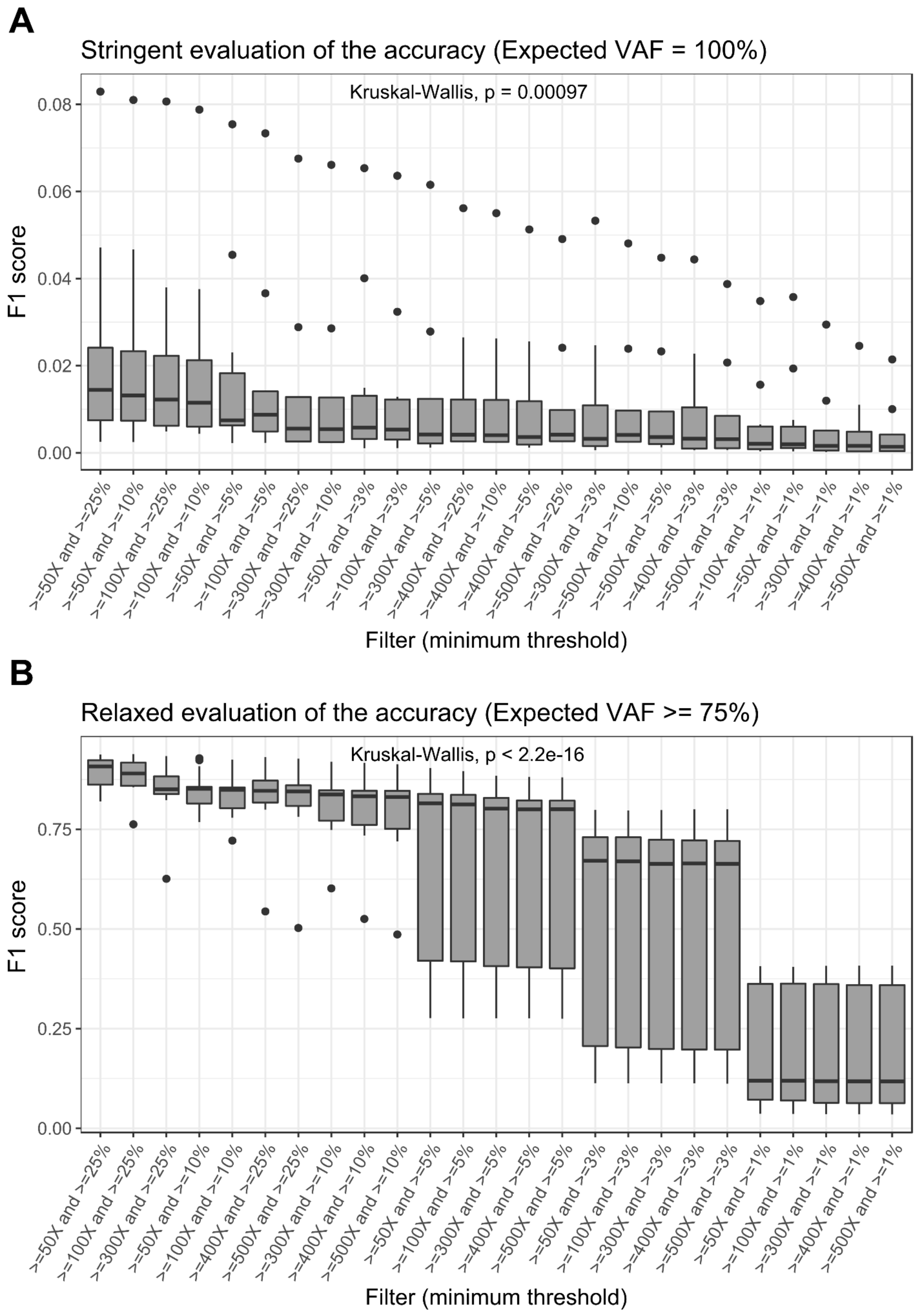


**Figure S1. Accuracy (F1 score) of SNV calling in positive controls as a function of depth and frequency filters.** A) F1 score of the SNV calling when the expected variant frequency is 100%. The Kruskal-Wallis test p-value (at the top of the panel) indicates the significance of the differences across the different sets of filters. The thresholds that define the filters are respectively the minimum coverage (X) and the minimum SNV frequency (%). B) F1 score of the SNV calling when the expected variant frequency is >=75%.

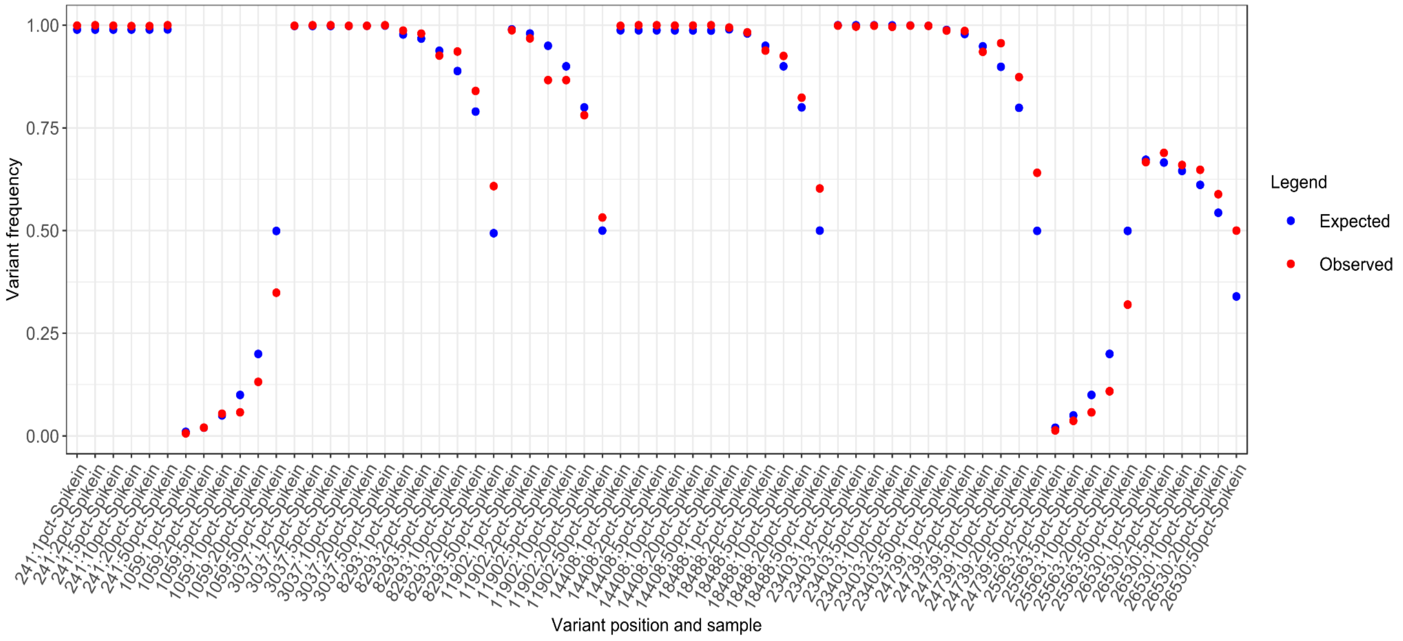


**Figure S2. Inferred SNV frequencies are in good agreement with expectation from 'spike-in' controls.** The expected variant frequency (VAF) are illustrated in blue while the observed/measured VAF from the sequencing data are illustrated in red. Nucleotide positions are listed in order across the SARS-CoV-2 gene along the x-axis.

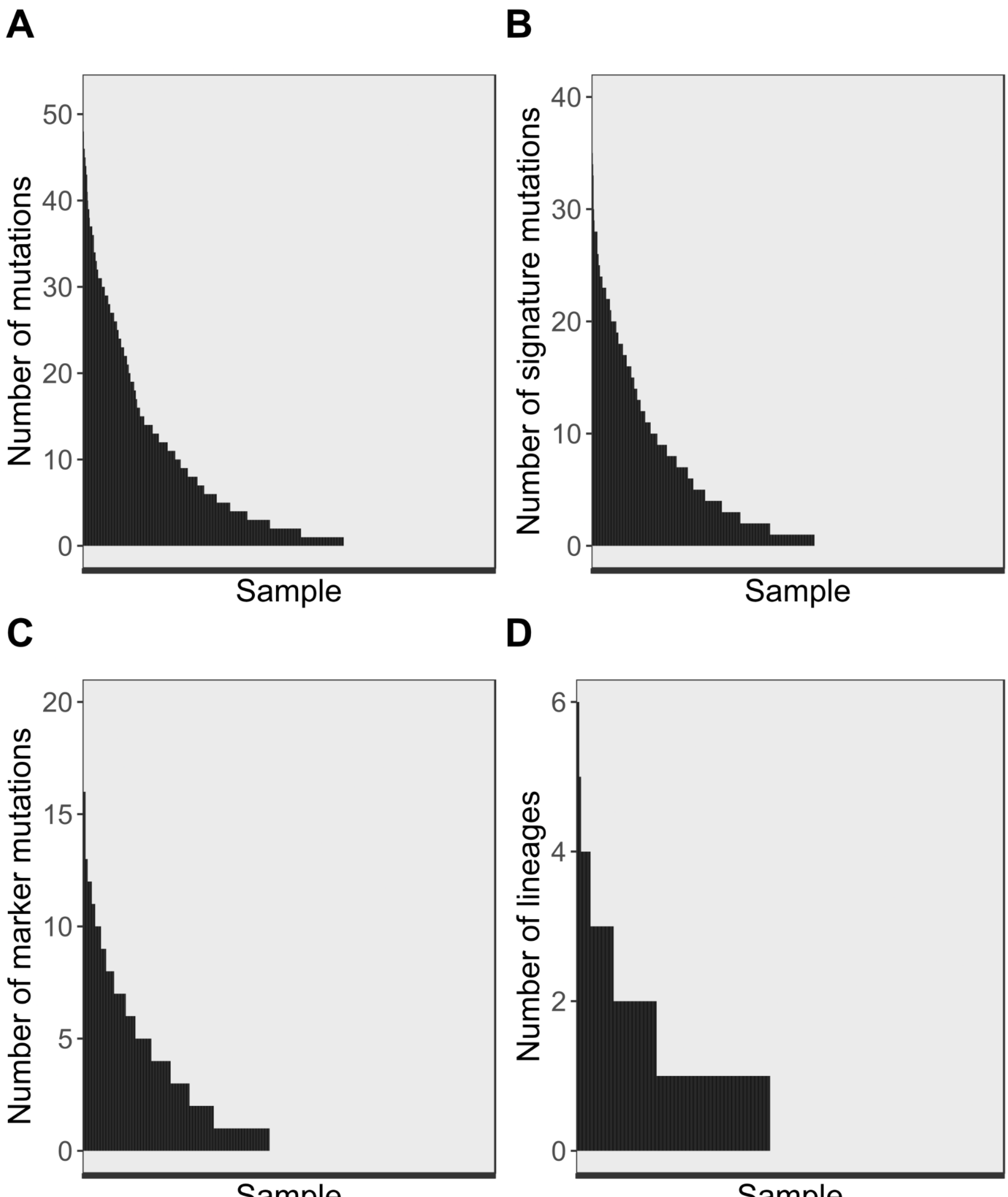


**Figure S3. Distribution of diversity metrics across samples.** A) Number of SNVs per sample (μ=6.87; s.d. = 10.1). B) Number of signatures mutations (μ=4.42; s.d. = 6.94). C) Number of marker mutations per sample (μ=2.01; s.d. = 3.22). D) Number of variant lineages inferred per sample (μ=0.79; s.d. = 1.14). Samples are ranked in descending order of the y-axis values.

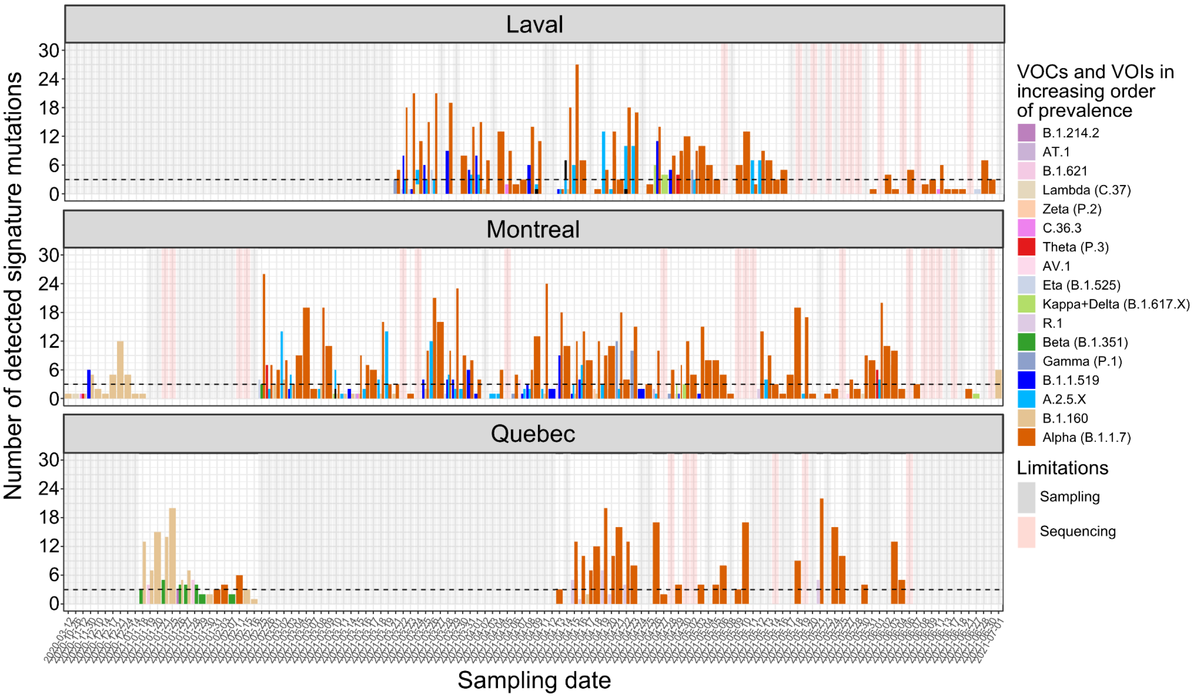


**Figure S4. Detection of variants of concern and variants of interest in Quebec wastewater samples.** Detected variants of concern (VOCs) and variants of interest (VOIs) are represented in different colors and ordered in increasing order of prevalence in the legend. Each panel represents a different city in Quebec. The black dotted line represents the minimum number of signature mutations required to infer the presence of a lineage in a WW sample, including at least one marker mutation. Lack of variant lineage detection can be explained by a lack of sampling (transparent grey), i.e. the absence of detections of a particular lineage due to the absence of samples, or missing detections in the sequencing data (transparent red), i.e. the absence of detections of a particular lineage although samples were collected during the corresponding period. The x-axis represents samples which were collected on a particular date, based on available data, and does not represent a linear time-scale.

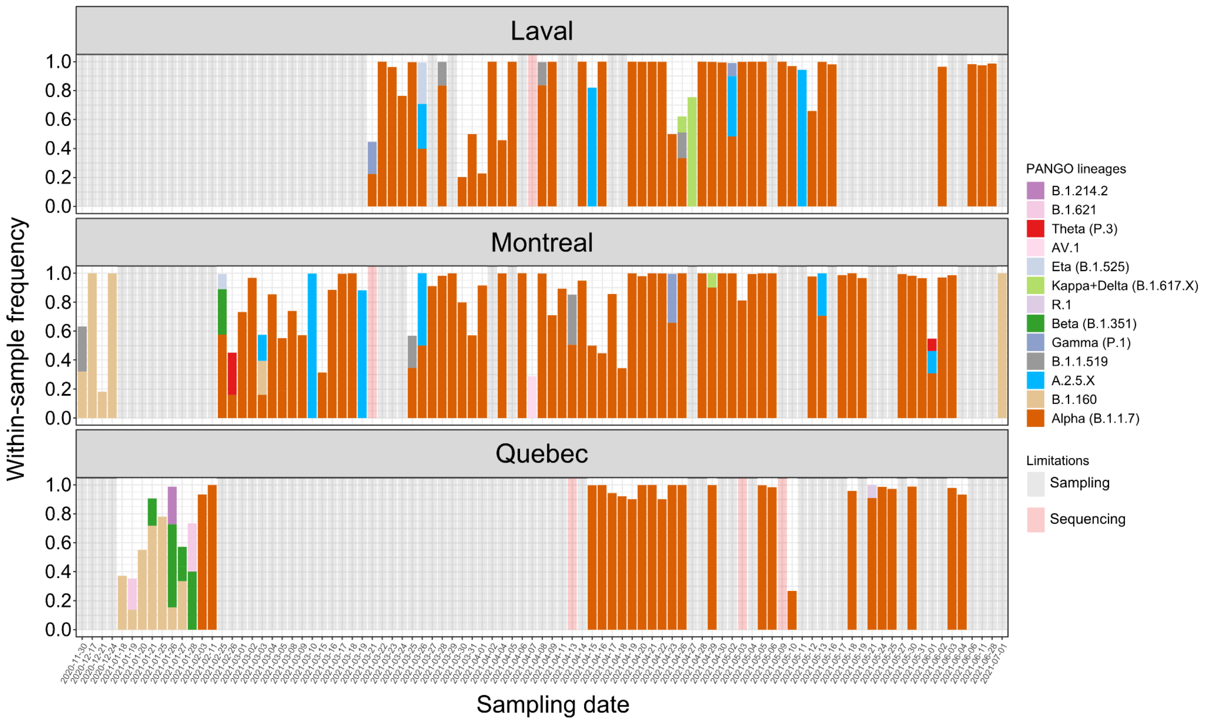


**Figure S5. Within-sample frequency of SARS-CoV-2 lineages.** Detected variants of concern (VOCs) and variants of interest (VOIs) are represented in different colors and ordered in increasing order of prevalence in the legend. Variant lineage frequencies were estimated with a constrained linear model (*t*-test $p < 0.05$). The x-axis represents samples which were collected on a particular date, based on available data, and does not represent a linear time-scale. Lack of variant lineage detection can be explained by a lack of sampling (transparent grey), i.e. the absence of detections of a particular lineage due to the absence of samples, or missing detections in the sequencing data (transparent red), i.e. the absence of detections of a particular lineage although samples were collected during the corresponding period.

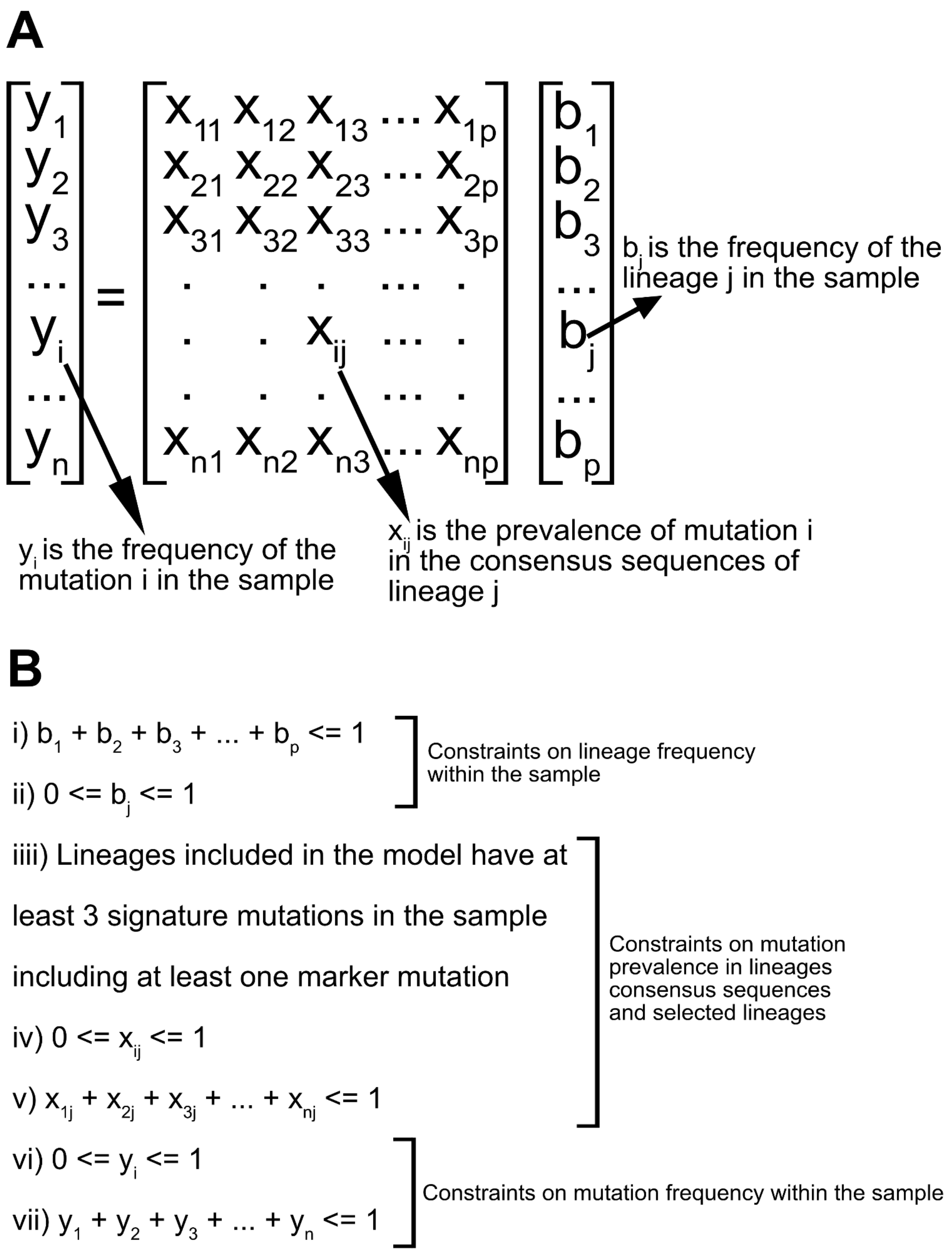


**Figure S6. Using constrained linear regression for estimating lineage within-sample frequency.** A) Linear regression model. The signature mutations' prevalence across the detected lineages has been fitted to the signature mutations frequency within each tested sample such that the regression coefficients are the lineage frequency when we applied the appropriate constraints. B) Constraints. The mutation and lineage frequency and prevalence have values between 0 and 1, and their respective sums are smaller or equal to 1. Only lineages with at least 3 detected signature mutations in the sample, including at least one marker mutation, are included in the model.

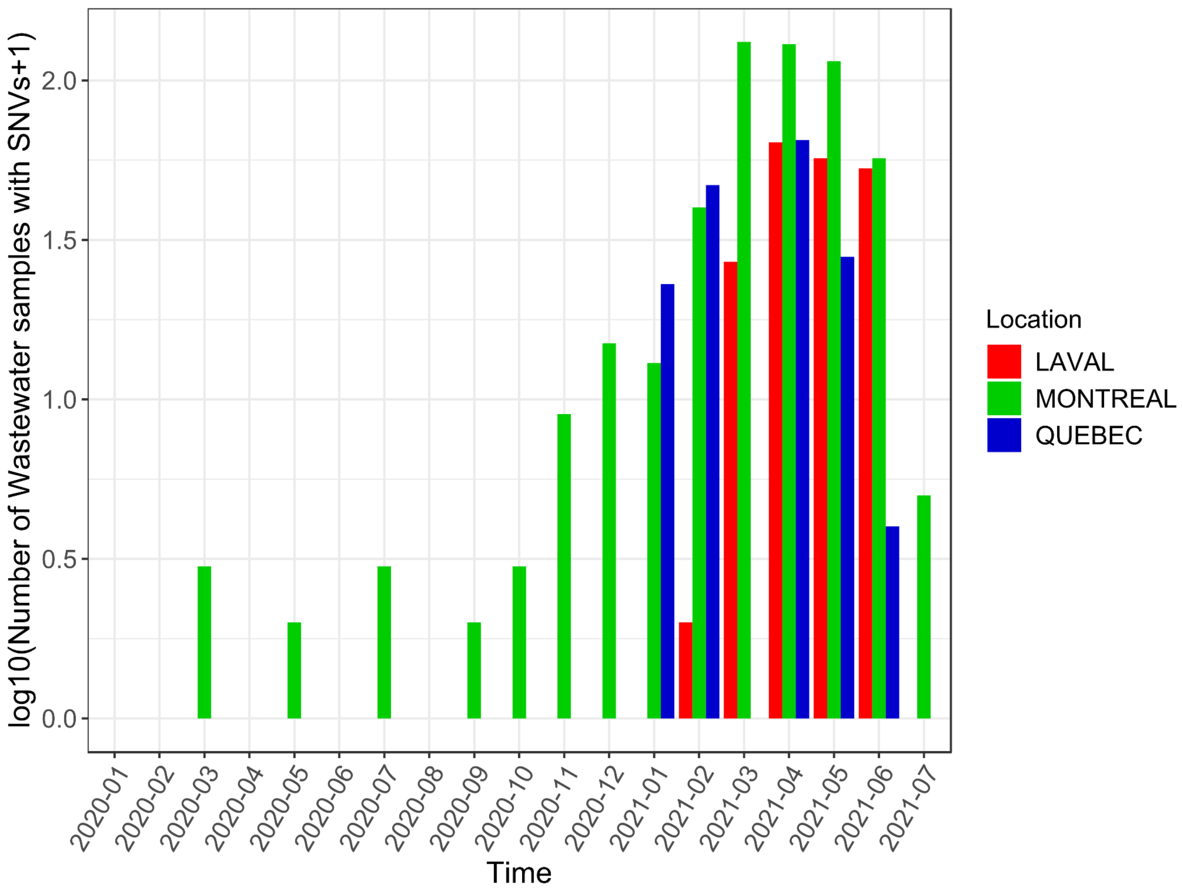


**Figure S7. Number of monthly wastewater samples with at least one SNV called across the locations under surveillance**.

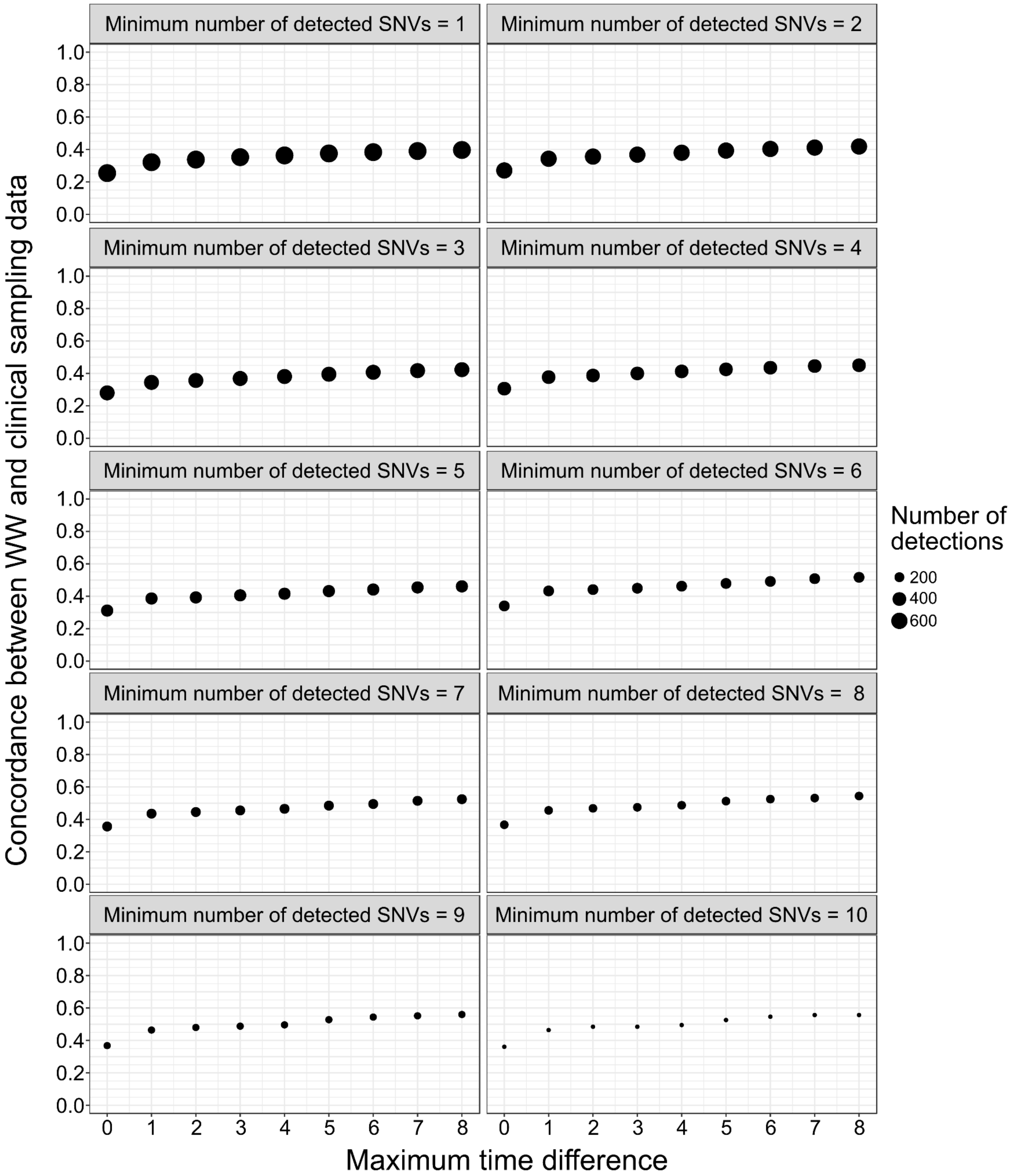


**Figure S8. Concordance of WW lineage detections with semi-random clinical data depending on the maximum time lag.** We defined concordance as the proportion of detections in the WW dataset that are also identified in the clinical sampling dataset. The size of the points represents the number of unique lineages detected using the set of filters. Each panel shows a different minimum number of signature SNVs required to define a PANGO lineage as present.

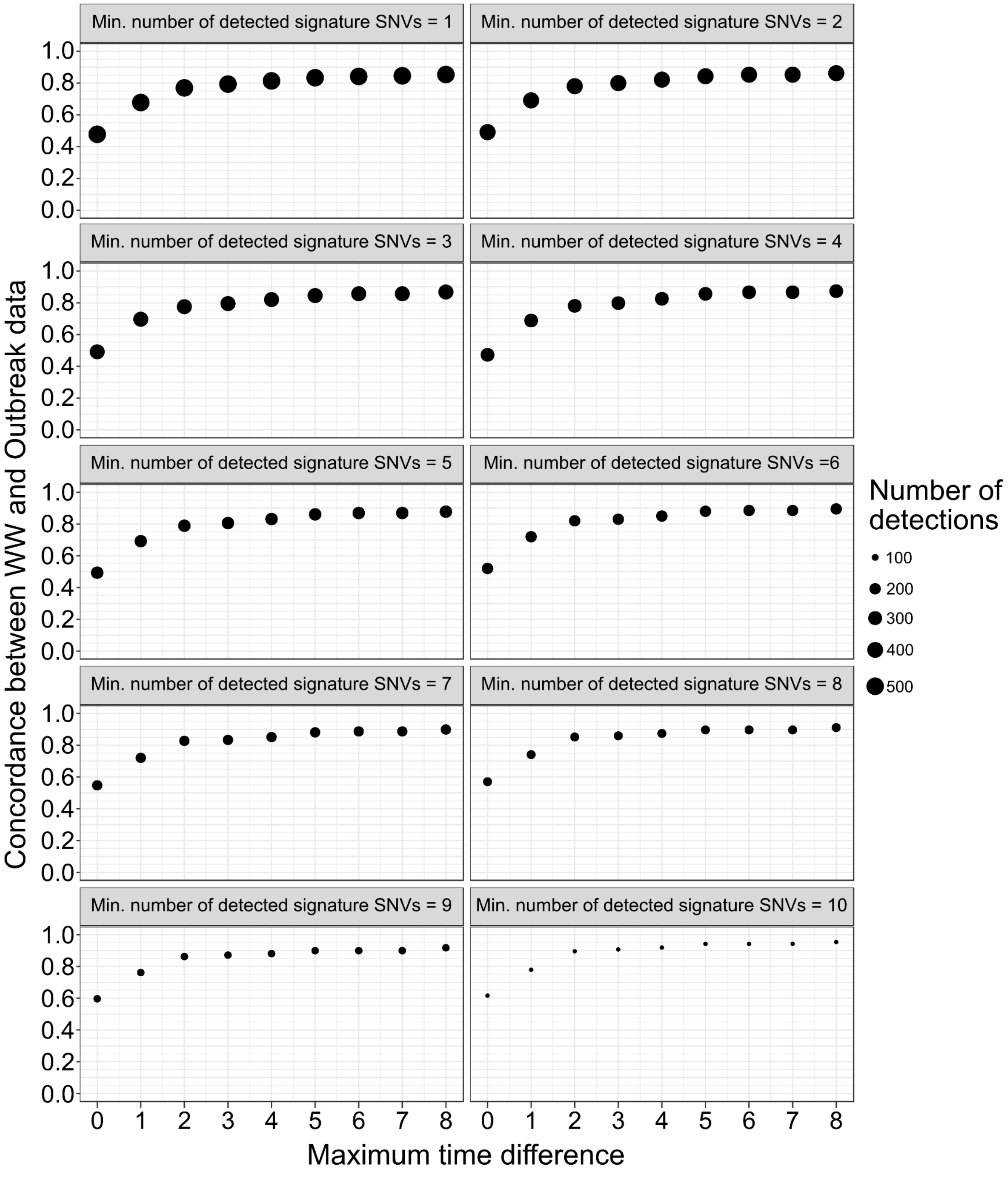


**Figure S9. Concordance of WW lineage detections with outbreak data depending on the maximum time lag.** We defined concordance as the proportion of detections in the WW dataset that are also identified in the outbreak dataset. The size of the points represents the number of unique lineages detected using the set of filters. Each panel shows a different minimum number of signature SNVs required to define a PANGO lineage as present.

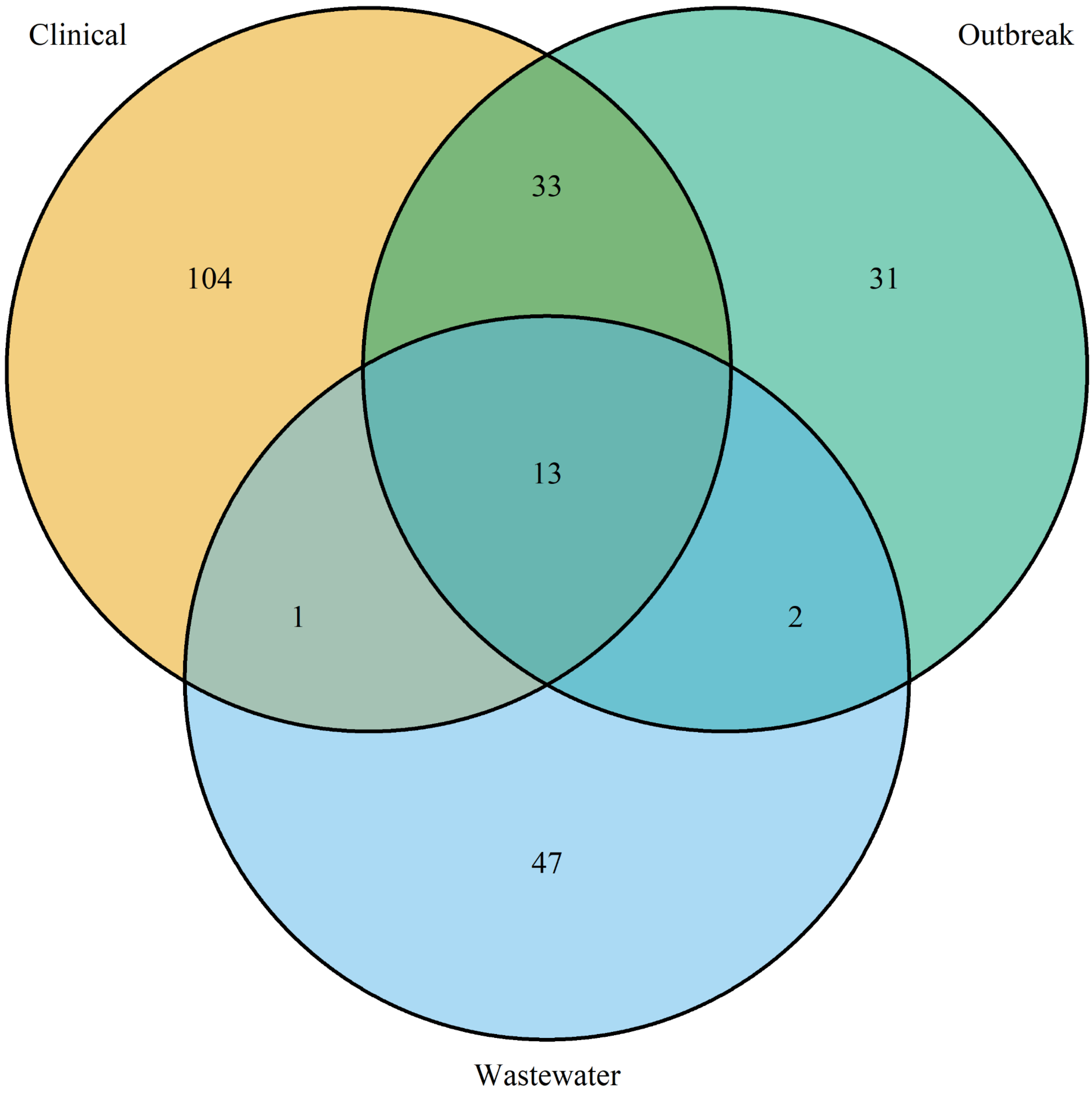


**Figure S10. Venn diagram of the variant lineages detected in WW, outbreak and clinical datasets.** Lineages detected in the clinical dataset are represented in gold, lineages detected in the outbreak dataset are represented in green and lineages detected in the wastewater dataset are represented in sky blue.

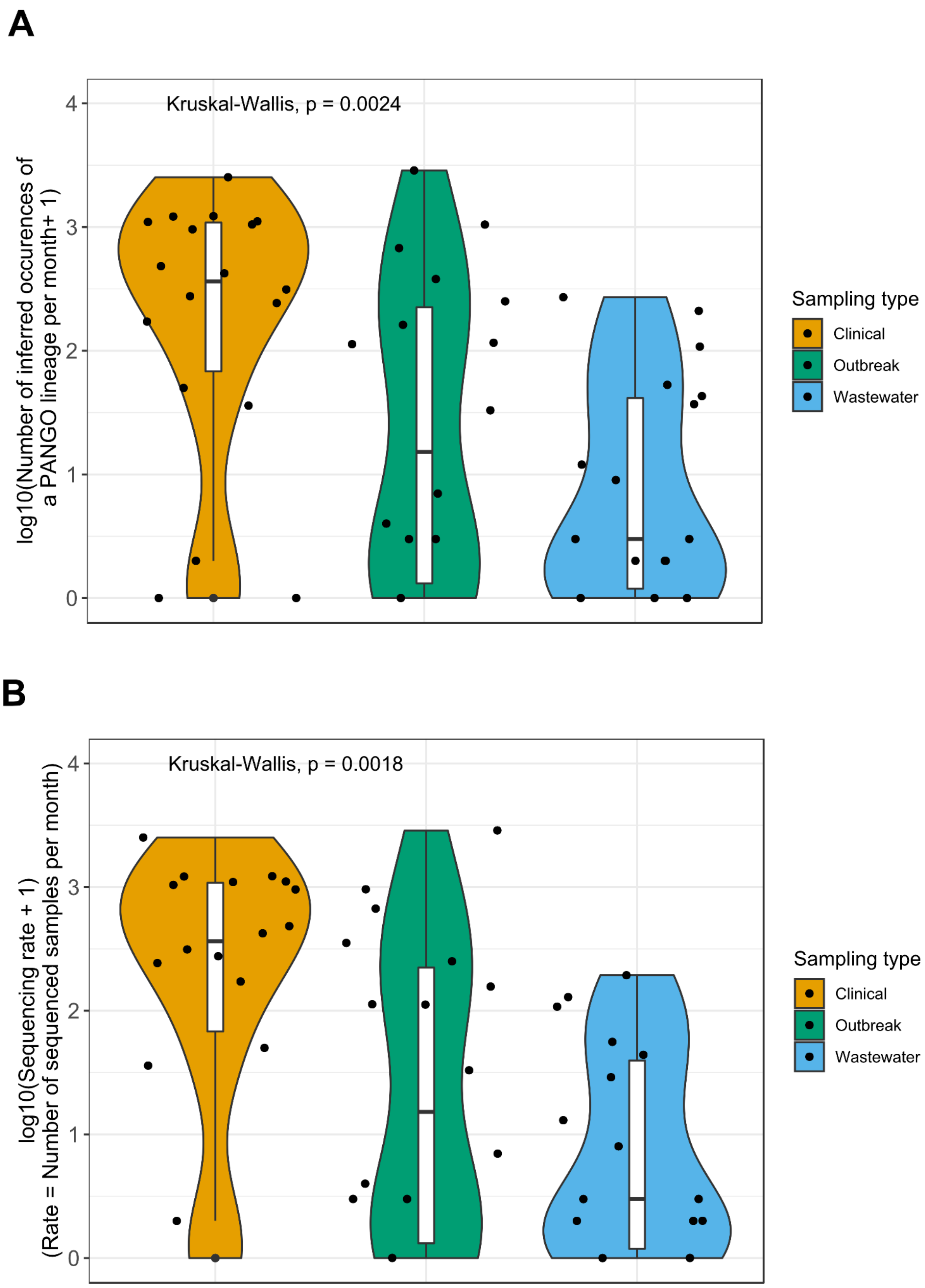


**Figure S11. Comparison of variant detection rates in WW and clinical sampling.** A) Distribution of the number of occurrences of lineages per month in clinical (gold), outbreak (green) and wastewater (sky blue) datasets. The Kruskal-Wallis *p* indicates the significance of the difference across the datasets. B) Distribution of the number of sequenced samples per month in clinical (gold), outbreak (green) and wastewater (sky blue) datasets.
